# Cohort-based association study of germline genetic variants with acute and chronic health complications of childhood cancer and its treatment: Genetic risks for childhood cancer complications Switzerland (GECCOS) study protocol

**DOI:** 10.1101/2021.01.30.21250679

**Authors:** Nicolas Waespe, Sven Strebel, Tiago Nava, Chakradhara Rao S. Uppugunduri, Denis Marino, Veneranda Mattiello, Maria Otth, Fabienne Gumy-Pause, André O. von Bueren, Frederic Baleydier, Luzius Mader, Adrian Spoerri, Claudia E. Kuehni, Marc Ansari

**Author notes:** **Corresponding author:** Prof. Dr. med. Marc Ansari, MD; Division of Pediatric Oncology and Hematology, Department of Women, Children, and Adolescents, University Hospital of Geneva, Rue Willy-Donzé 6; Bureau 5-507; 1211 Genève, Switzerland; F.

## Abstract

**Background:** Childhood cancer and its treatment may lead to many acute and chronic health complications. Related impairment in quality of life, excess in deaths, and accumulated health care costs are relevant. There is a wide inter-individual variability in the type and severity of health complications. Genetic variations are suggested to contribute to individual susceptibility. So far, only few genetic variants have been used to risk-stratify treatment and follow-up care. This study platform aims to identify germline genetic variants associated with acute and late complications of childhood cancer.

**Methods:** The Genetic Risks for Childhood Cancer Complications Switzerland (GECCOS) study is a nationwide cohort study. It includes patients and survivors who were diagnosed with childhood cancers or Langerhans cell histiocytosis before age 21 years, were registered in the Swiss Childhood Cancer Registry (SCCR) since 1976 and have consented to the Pediatric Biobank for Research in Hematology and Oncology (BaHOP), Geneva, host of the Germline DNA Biobank Switzerland for Childhood Cancer and Blood Disorders (BISKIDS). BISKIDS is a national biobank for the collection of germline DNA in childhood cancer patients and survivors.

GECCOS uses demographic and clinical data from the SCCR and the associated Swiss Childhood Cancer Survivor Study (SCCSS), which contains health-related data of survivors. Phenotypic data consist of objective measurements, health conditions diagnosed by physicians, second primary neoplasms, self-reported and health-related information from participants. Germline genetic samples and sequencing data have been collected in BISKIDS. We will perform gene panel sequencing, whole-exome sequencing, or whole-genome sequencing depending on the research questions. We will perform association analyses to identify genetic variants associated with specified health conditions. We will use clustering and machine-learning techniques and assess multiple health conditions in different models.

**Discussion:** GECCOS will serve as an overarching platform to enable genotype-phenotype association analyses on complications associated with childhood cancer and its treatments. Knowledge of germline genetic variants associated with childhood cancer-associated health conditions will help to further individualize cancer treatment and follow-up care, potentially resulting in improved efficacy and reduced side effects, for personalized cancer care.

**Trial registration:** Clinicaltrials.gov: NCT04702321

## MAIN MANUSCRIPT

### 1. Background

Childhood cancers have become curable in ≥85% of patients in developed countries.[1] Current treatment protocols are multimodal with varying combinations of surgery, chemotherapy, radiation, hematopoietic stem cell transplantation (HSCT), and immunotherapies. The price of increased survival is a wide range of acute and chronic health conditions. Cancer treatments are associated with acute complications such as transient nausea and vomiting, mucositis, and fatigue but also pneumonitis, cardiomyopathy, encephalitis, and life-threatening infections.[2] While many of these conditions are potentially reversible, some are not and become chronic or develop over time like cardiac, pulmonary, auditory, endocrine, reproductive, and neurocognitive health conditions, and second primary neoplasms (SPNs).[3] A recent publication found a cumulative incidence of severe chronic health conditions of 96% in childhood cancer survivors aged 50 years. The number of severe chronic health conditions was 2 times higher in survivors compared to matched community controls.[4] Mortality is significantly increased in survivors compared to the general population.[5] Recurrence and SPNs are the leading causes of death in the first 2 decades after cancer diagnosis followed by diseases of the cardiovascular and respiratory systems thereafter.[5, 6] Because of their young age at diagnosis, survivors have decades of life time ahead. The burden of chronic conditions, related impairment in quality of life, excess in deaths, and accumulated health care costs is therefore of great relevance for them.[7]

Only few genetic variants modifying the risk of acute and chronic toxicities in children with cancer have so far led to personalized treatment protocols or follow-up care.[8, 9] An example is 6-mercaptopurine, where dosing is routinely adapted in contemporary treatment protocols for patients with thiopurine methyltransferase (*TPMT*) variants which increase the risk of acute hematological toxicity in the treatment of childhood acute lymphoblastic leukemia (ALL).[10] Other genetic markers have been associated with various outcomes but so far not implemented in treatment protocols, such as nudix hydrolase 15 *(NUDT15)* with hematological toxicity after purine analogue treatment,[11, 12] dihydrofolate reductase *(DHFR)* with overall survival in some subtypes of ALL,[13, 14] and glutathione S transferase genes with various outcomes after hematopoietic stem cell transplantation.[15] Also, the effect of genetic variation on late toxicities, chronic health conditions arising after the end of childhood cancer treatment, has been investigated.[16, 17] Different clinical outcomes such as hearing loss,[18–21] cardiomyopathy,[22, 23] metabolic syndrome,[24, 25] and gonadal impairment have been studied.[26, 27] Exome-wide (EWAS) and genome-wide association studies (GWAS) have led to the identification of genetic variants associated with drug toxicities (e.g. osteonecrosis in children with ALL [*BMP7* and *PROX1-AS1*], asparaginase hypersensitivity [*GRIA1*], or vincristine-associated peripheral neuropathy in children with ALL [*CEP72*]),[28–31] metabolic syndrome,[24] sinusoidal obstruction syndrome after HSCT,[32] and hearing loss.[18] Acylphosphatase 2 (*ACYP2*) was associated with hearing loss after platinum treatment exposure in several independent datasets but failed to be replicated in a recent large candidate gene analysis while a variant in solute carrier family 22, member 2 (*SLC22A2*) was found to be associated with mild hearing loss.[20, 33] Genetic modifiers have been implicated for the development of SPNs including breast cancer,[34] CNS tumors,[35] and leukemia.[36] For other SPNs (such as thyroid cancer), no data is available.[37] To address the contribution of genetic risk variants in the development of late toxicities, large cancer survivor studies such as the Childhood Cancer Survivor Study (CCSS) in the US are collecting DNA systematically to conduct genotype-phenotype analyses.[36] Most studies on genetic risk variants used a candidate-gene approach, had a small sample size of less than 200 participants, heterogenous cohorts with various treatment exposures, and inconsistent outcome assessments.[17] Many health conditions have not been investigated, like renal insufficiency, pulmonary, and ocular complications. It is likely that several distinct pathways and their corresponding gene variants are involved in the development of complex phenotypes like pulmonary dysfunction. Therefore, many candidate gene variants and treatment exposures need to be considered. This is possible with hypothesis-free exome-wide or genome-wide association studies (EWAS or GWAS). New analytical approaches which combine clinical, pharmacological, and genetic data into integrative models have been developed and are showing promising results.[38, 39] Network analyses, machine learning, and clustering methods might help to understand the impact of genetic variations on complex phenotypes with biological pathways and their corresponding genes. The GECCOS study will enable large genotype-phenotype association studies in childhood cancer patients and survivors in Switzerland and international collaborations. Selected organ dysfunctions and second primary neoplasms will be studied in sub-projects, and *in silico* and *in vitro* methods will be used to further explore mechanisms associated with genetic variation and outcomes.

### 2. Study objectives

The main objectives of the GECCOS study are

i. to identify genetic variants associated with health conditions in childhood cancer and its treatment using genotype-phenotype association methods;
ii. to evaluate the biological function of genetic variants associated with health conditions through *in silic*o and *in vitro* methods; and
iii. to identify genetic variants associated with multiple health conditions in childhood cancer using models integrating multiple outcomes.

The secondary objectives are

a. to create a common framework for sub-studies using genotype-phenotype associations with germline genetic material and data of childhood cancer patients and survivors;
b. to feed germline genetic data generated in sub-studies into a biobank database for future research and create a growing repository of genetic sequencing data; and
c. to facilitate research using a common structure that can be used for collaborations.

### 3. Methods

#### 3.1. Study design

The GECCOS study is a nationwide cohort study in collaboration of the Pediatric Biobank for Research in Hematology and Oncology (BaHOP) Geneva, Switzerland, and the Institute for Social and Preventive Medicine at the University of Bern. The Geneva Cantonal Commission for Research Ethics has approved the GECCOS study (approval 2020-01723), and the BaHOP biobank (approval PB_2017-00533).

#### 3.2. Data sources

The GECCOS study uses genetic data and material from BaHOP. Clinical information is collected from (i) the Swiss Childhood Cancer Registry (SCCR) and (ii) the Swiss Childhood Cancer Survivor Study (SCCSS; **Figure 1**). The nationwide germline DNA biobank for childhood cancer and blood disorders (BISKIDS) was established in May 2019 within the Biobank for Research in Hematology and Oncology (BaHOP) with support of all nine pediatric oncology centers caring for childhood cancer patients in Switzerland. Return of results and relevant incidental outcomes to the patient is defined in the BaHOP regulations with oversight of a genetic advisory board. BISKIDS collects germline DNA samples, extracts and stores genomic DNA and genetic data of childhood cancer patients as well as survivors in Switzerland.

**Figure 1.**
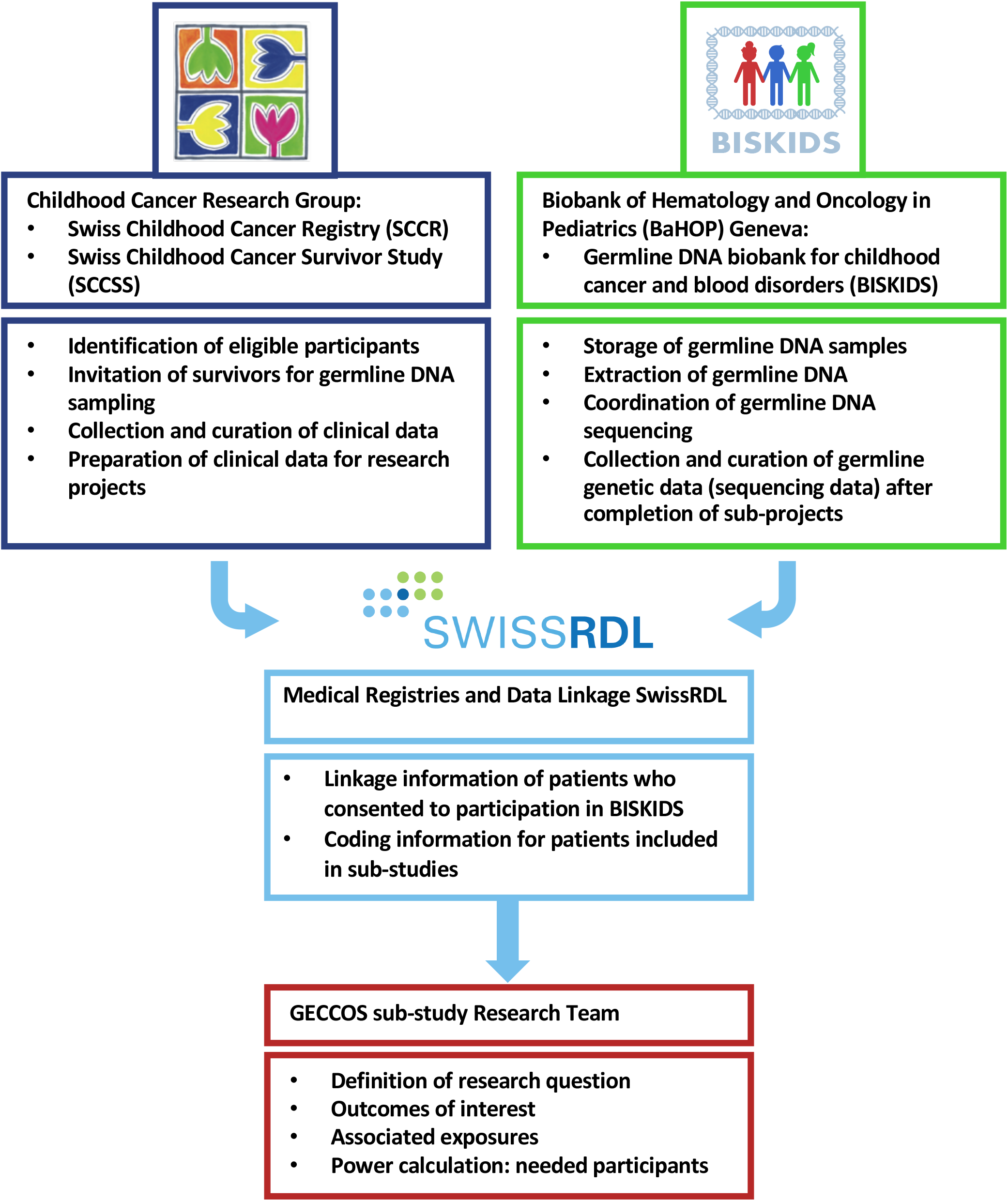
Responsible teams in the GECCOS study for germline genetic association with health conditions in childhood cancer patients and survivors. Legend: BaHOP, Biobank for Hematology and Oncology in Pediatrics (BaHOP); BISKIDS, Germline DNA Biobank for childhood cancer and blood disorders; DNA, deoxyribonucleic acid; SCCR, Swiss Childhood Cancer Registry; SCCSS, Swiss Childhood Cancer Survivor Study

The SCCR collects information on childhood cancer patients diagnosed in Switzerland.[40] Children and adolescents aged <21 years with a primary cancer diagnosis according to the International Classification of Childhood Cancer, third edition (ICCC-3), and Langerhans cell histiocytosis were registered since 1976 in the SCCR. The dataset is managed by the Childhood Cancer Research Group at the Institute of Social and Preventive Medicine, University of Bern. It has a completeness of coverage of childhood cancer patients in Switzerland aged up to 15 years of >85% since 1985 and >95% since 1995.[41] As of December 2019, 13,029 patients were registered in the SCCR, of which 9,306 (69%) were still alive and potentially eligible for participation in GECCOS. Mandated by a national cancer registration law in Switzerland enacted on January 1^st^, 2020, registration of new cancer patients is performed by the federal government from that date onwards.[42] The Institute of Social and Preventive Medicine of the University of Bern was commissioned to perform patient registrations and data collection, and the Childhood Cancer Research Group will continue research activities on these datasets. The GECCOS project will seek to include patients diagnosed with neoplasms during childhood after January 1^st^, 2020, and continue collaboration with the Childhood Cancer Research Group at the University of Bern. The Swiss Childhood Cancer Survivor Study (SCCSS, clinicaltrials.gov: NCT03297034) is a population-based, long-term cohort study of all childhood cancer patients who were registered in the SCCR, resident in Switzerland at diagnosis, and survived ≥5 years after initial cancer diagnosis.[43] The SCCSS collects questionnaire-based information from survivors, and data on chronic health conditions after childhood cancer from survivorship clinics and hospital records (e.g. audiograms, and lung function tests) which will be available for GECCOS.

#### 3.3. Study population

Eligible in the participation in the GECCOS study are persons who:

1. are registered in the Swiss Childhood Cancer Registry; and
2. were diagnosed with a neoplasm according to the ICCC-3, or Langerhans cell histiocytosis (LCH) before age 21 years; and
3. have consented themselves or through their legal representative to the BaHOP (host biobank for BISKIDS).

Inclusion criteria for sub-projects focusing on specific health conditions will be defined (**Supplementary Table 1 and 2** for established sub-projects). Within sub-projects, we will sample patients and survivors according to risk exposure (for a cohort design), or according to the outcome of interest (for a case-control or case-cohort design). We will identify eligible participants from the SCCR and SCCSS.

**Table 1.** Summary information on main covariates and exposures of interest used in the genotype-phenotype association analyses.

| **Covariate type** | **Covariates** | **Unit** |
| --- | --- | --- |
| **Demographic information** | Sex | Male/ female/ other |
|  | Birthdate | Month/ year |
|  | Country of origin | Country name |
| **Socio-economic status** | Highest education patient and parents | 10-unit scale |
|  | Income patient and/ or parents | Monthly net income (Swiss francs) |
| **First primary neoplasm information** | Age at diagnosis | Years |
|  | Date of diagnosis | Month/ year |
|  | Type of diagnosis | ICCC3 code; ICDO3 morphology, topography, behavior code |
|  | Laterality | Left/ right/ bilateral/ medial/ not applicable |
|  | Relapse date | Month/ year |
|  | Relapse type | Local/ distant/ systemic/ other |
|  | Relapse location | Organ and morphology |
| **Treatment information** | Treatment protocol | Name of protocol, arm, randomization group |
| **Chemotherapy** | Separately per antineoplastic agent: Cumulative dose estimated using treatment protocols or if available calculated using extracted data from medical records; | mg/m<sup>2</sup>, or appropriate metric; cycles (n); dose per cycle (mg/m<sup>2</sup>) |
|  | Start date | Month/ year |
| **Radiotherapy** | Separately per radiation field: Cumulative dose estimated on an intention to treat basis using radiotherapy field descriptions, or calculated from radiotherapy protocols | Mean and maximal estimated dose per organ (Gy); fractions (n); dose per fraction (Gy) |
|  | Radiation type | Photon, proton, brachytherapy, stereotactic radiation, other |
|  | Radiation field | Description of radiation field (e.g. mantle field, whole lung irradiation, total body irradiation) |
|  | Laterality | Left/ right/ bilateral/ medial/ not applicable |
|  | Start date | Month/ year |
|  | Concomitant chemotherapy | Antineoplastic agent and dose (mg/m2) |
| **Surgery** | Location | Organ, site, and description of intervention |
|  | Laterality | Left/ right/ bilateral/ medial/ not applicable |
|  | Date | Month/ year |
| **Hematopoietic stem cell transplantation** | Type | Allogeneic, autologous, other |
|  | Donor type | Matched sibling, matched other relative, haploidentical relative, matched unrelated, mismatched unrelated, other |
|  | Donor graft source | Bone marrow, peripheral stem cells (apheresis), umbilical cord blood, other |
|  | Matching degree | Number of HLA loci matched of total assessed HLA loci |
|  | Conditioning regimen | Name and treatment details |
|  | Total body irradiation | Yes (including dose in Gy), no |
|  | Date of stem cell transfer | Month/ year |
|  | Acute complications | Sinusoidal obstruction syndrome, infection, acute GvHD (with grading), others |
|  | Chronic complications | Chronic GvHD (with grading) |
| **Follow-up information** | Last vital status | Alive, dead, unknown |
|  | Date of last vital status | Month/ year |
|  | Last follow-up information from clinical site | Month/ year |

Legend: GvHD, graft versus host disease; Gy, Gray; HLA, human leukocyte antigen; ICC3, International Classification of Childhood Cancer, third edition; ICDO3, International Classification of Diseases for Oncology, third edition; mg/m2, milligrams per square meter; n, number.

**Table 2.** Summary information on the genetic information used in the genotype-phenotype association analyses.

| **Information** | **Data** | **Format** |
| --- | --- | --- |
| **Germline genetic sequencing data** | Raw sequencing data | FASTQ or BAM files |
|  | Identified genetic variants | VCF files |
|  | Documentation of sequencing procedure and analyses performed to allow trackability of downstream analyses | CSV or TXT files to allow long-term readability |
|  | Quality measures | Read depth, score to evaluate correct variant calling, etc. |
| **Underlying genetic condition** | Heritable underlying condition | Description of underlying condition |
|  | Gene name of affected gene | HGVS name |
|  | Identified causal variant | HGVS notation: genomic location, coding DNA, and expected effect on protein; rs number if available |

Legend: BAM, compressed binary file used to represent aligned sequences; CSV, comma separated values; DNA, deoxyribonucleic acid; FASTQ, text file containing the sequence data from the clusters that pass filter on a flow cell; HGVS, Human Genome Variation Society; rs, reference single nucleotide polymorphism identifier; TXT, standard unformatted text document; VCF, variant call format.

#### 3.4. Outcomes: health conditions and second primary neoplasms

We will assess health conditions in childhood cancer patients and survivors by collecting data on organ function such as pulmonary functions tests for lung conditions, or audiograms for hearing loss. For outcomes that cannot be adequately measured (e.g. tinnitus), we will use information from self-assessment questionnaires. We will use the International Agency for Research on Cancer (IARC) criteria for coding SPNs.[44] In brief, we will include neoplasms according to the ICCC-3 classification which originated in different tissues or had a different morphology than the first primary neoplasm. We will not classify progression, transformation, metastasis, and relapse of first primary neoplasm as SPN.

#### 3.5. Covariates

For specific sub-studies and analyses, we will extract data on relevant covariates that might influence the outcomes of interest from the SCCR and SCCSS. We will extract demographic, socio-economic, first primary neoplasm, treatment, and follow-up information. For treatment information, we will estimate cumulative doses of chemotherapies using individual treatment protocols, or calculate effective treatment doses from medical records, if available. We will estimate exposure to radiotherapy using radiotherapy field descriptions,[45–47] or calculate organ-specific exposure from effective administered radiotherapy documentation (**Table 1**).

#### 3.6. Selection of participants

We will identify participants eligible for specific sub-studies with defined in- and exclusion criteria. We will use information in the SCCR and SCCSS and assess availability of corresponding germline DNA samples or sequencing data from previously sequenced participants in BISKIDS (**Figure 2**). If clinical data is available for a sufficient number of participants but further genetic samples are needed, we will invite potential participants to contribute germline DNA samples to BISKIDS for research. For collection of biological material within BISKIDS, we will use two pathways: (i) invitations to participate are sent out by the Childhood Cancer Research group at the University of Bern, consisting of germline DNA collection kits (predominantly using saliva samples or buccal swabs) with information on the biobanking project and associated research, and informed consents to the participant’s home; (ii) participants are invited by healthcare staff in hospitals caring for childhood cancer patients and survivors. These potential participants and their legal representatives are informed of the project and written consent and germline DNA are collected during a medical visit already planned for their treatment or follow-up. All participants consent to have their germline DNA stored in the BISKIDS section of the BaHOP biobank and their health-related data to be used for genotype-phenotype association studies. All specific GECCOS sub-studies will be reviewed and approved by a national scientific committee and submitted to the responsible ethics committee as amendment to the main GECCOS protocol, insofar as the applicable law requires authorization.

**Figure 2.**
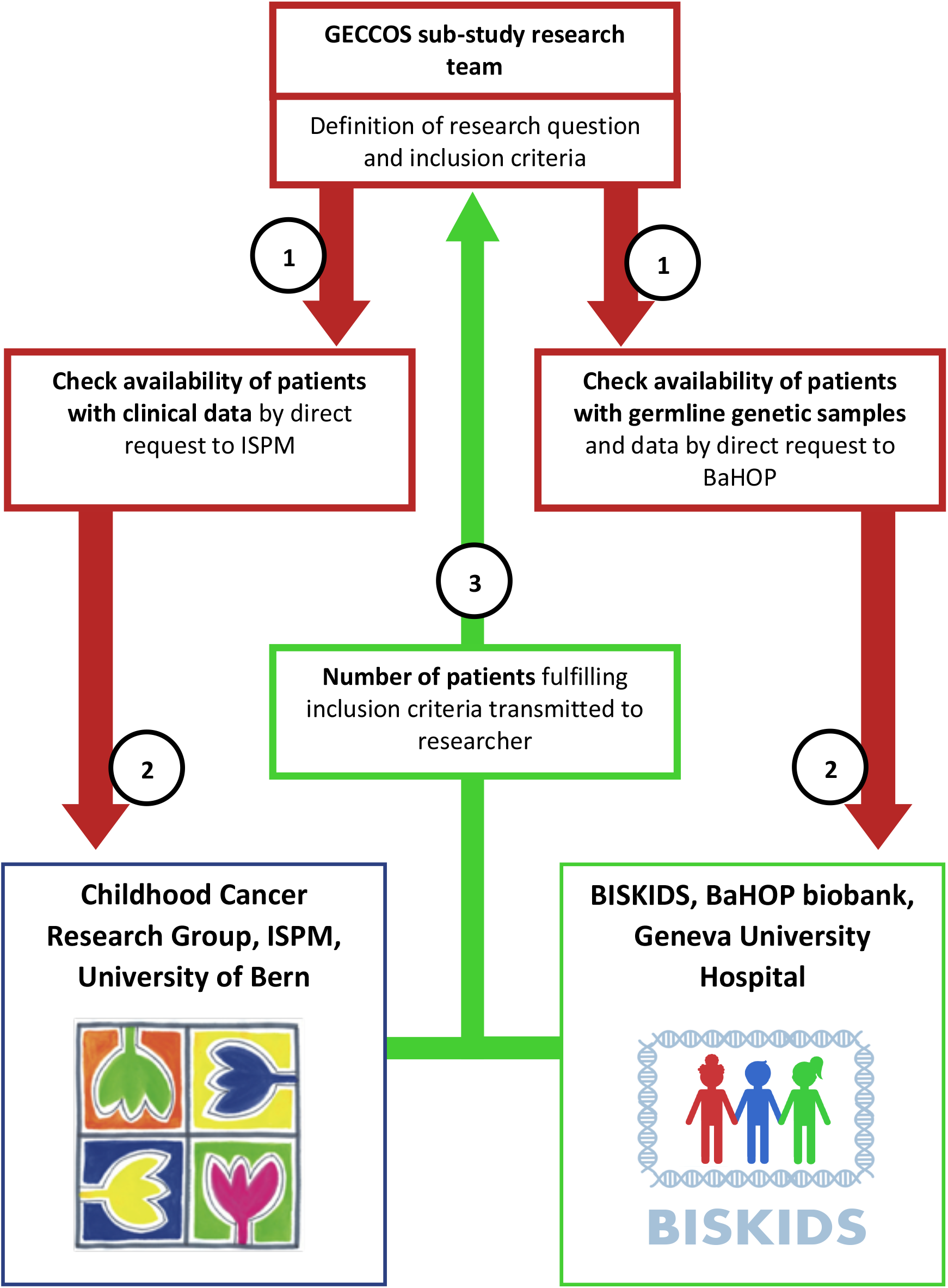
Flow diagram of identification of eligible participants for a specific sub-project: (1) The eligibility criteria, as defined by the researchers, will be transmitted to the Childhood Cancer Research Group at the ISPM, University of Bern, and the BISKIDS biobank at the Geneva University Hospital; (2) the number of eligible participants is compiled by secured data exchange from the BISKIDS project and the Childhood Cancer Research Group; (3) The number of eligible participants will be transmitted to the researcher to assess feasibility of a sub-project.

#### 3.7. Data linkage

Information allowing to link genotype data from the BISKIDS collection with phenotype data from the SCCR and SCCSS is securely stored in a separate trust center database managed by a third party (SwissRDL – Swiss Medical Registries and Data Linkage, ISPM, University of Bern). This procedure allows to separate identifying information from clinical and genetic information. We use a web-based secured and personalized access point (currently RedCap 9.5, Vanderbilt University, Nashville, TN, USA). The key located at the trust center is used to merge the clinical dataset with the germline genetic information without releasing identifying information to the GECCOS research team or to the BaHOP biobank. The research dataset will only contain a unique study-specific patient-identifier, without any identifying information. Study-specific identifiers will be securely stored in the trust center database to assure traceability of datasets (**Figure 3**).

**Figure 3.**
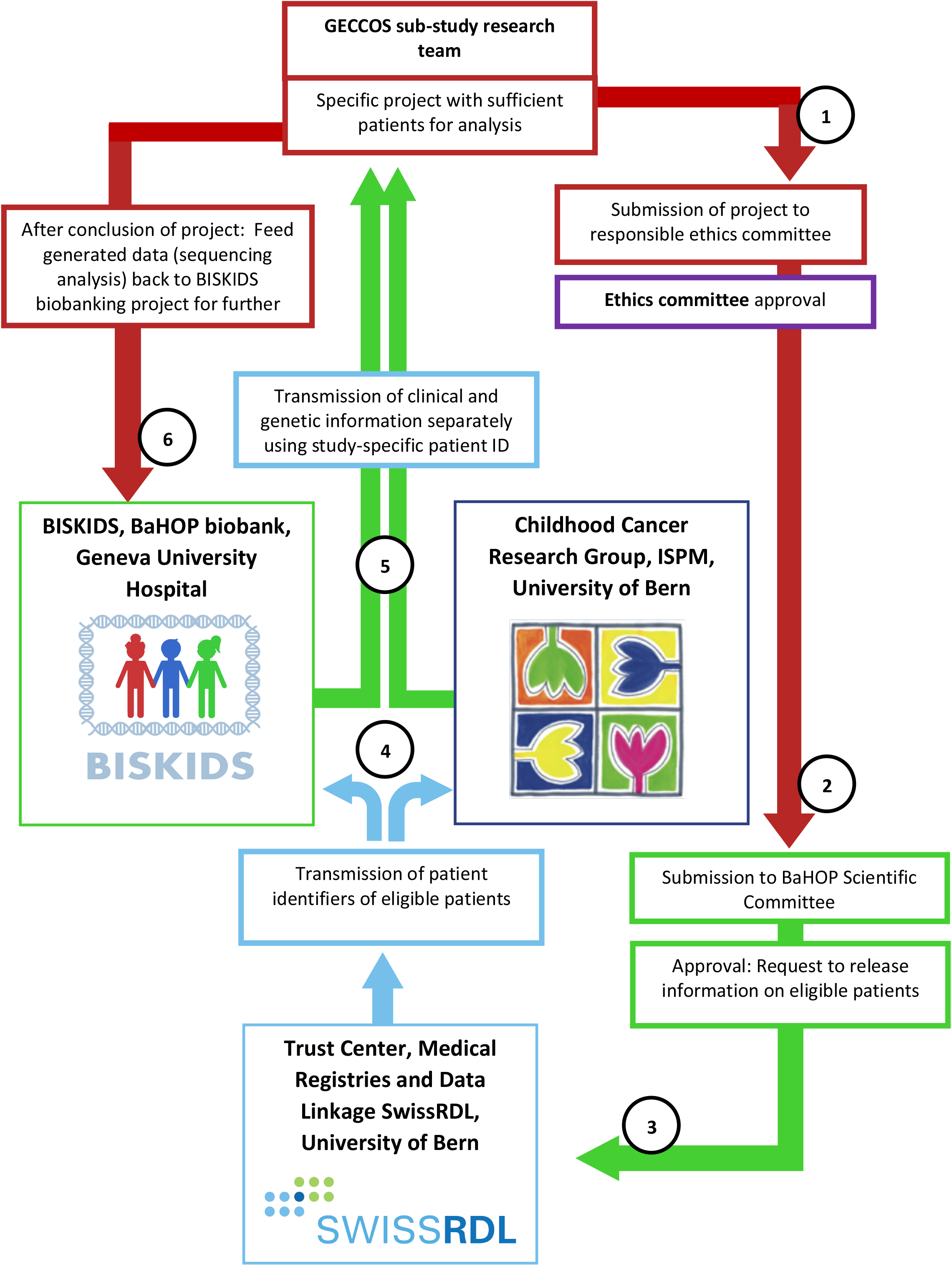
Flow diagram of release of information from the different resources. (1) Submission of sub-project to the responsible ethics committee, either as amendment to the main protocol of GECCOS or as a separate project; (2) Submission of sub-project to the scientific committee of the BaHOP, host biobank for the BISKIDS section at the Geneva University Hospital; (3) Release of linking information from SwissRDL; (4) Transmission of BISKIDS and SCCR identifiers for included patients with a newly generated study-specific patient-identifier to BISKIDS and the ISPM respectively, to release variables used for the study; (5) Transfer of data for included participants with a data transfer agreement to the researcher with study-specific identifier; (6) After conclusion of the project: Storage of acquired germline genetic data in the BISKIDS biobanking database at the Geneva University Hospital for future research projects.

#### 3.8. Data and sample handling

Clinical data will be transferred in standard data formats. Analysis datasets will not include identifying information. Management of germline DNA samples (e.g. saliva or buccal swabs), DNA extraction, aliquoting, and storage procedures are clearly defined in the BaHOP biobank regulation. For germline DNA sequencing in the GECCOS study, one of the DNA aliquots will be sent to Campus Biotech, Geneva, a sequencing facility. For genotype-phenotype analyses, we will collaborate with the Swiss Institute of Bioinformatics, Switzerland. Sequencing data and relevant clinical outcome data will be shared in a secured and encrypted way between the sequencing facility, the Swiss Institute of Bioinformatics for analysis, and the research platform for pediatric oncology and hematology in Geneva. Data will be stored and made available through a harmonized nationwide network to support computational biomedical research and clinical bioinformatics.[48]

#### 3.9. Sequencing analyses

We will use genetic information (**Table 2**) from different genomic sequencing methods: (I) gene panel sequencing adapted to specific research questions (using TruSeq DNA PCR free library preparation kit, Illumina, San Diego, USA); (II) whole-exome sequencing (using Illumina HiSeq4000 or NovaSeq 6000 platform with a mean read depth of at least 70x); and (III) whole-genome sequencing (with a mean read depth of at least 30x), depending on the research question. We will use workflows for genotyping implemented in the Genome Analysis Toolkit (GATK)[49] and adapted to the aim of the study for (1) sequence generation; (2) sequence alignment; (3) variant calling; (4) variant filtering; and (5) variant annotation.[50] We will also include data on read quality control. We will perform analyses with any of the following: (i) a candidate gene approach with filtered variants in pre-selected genes based on scientific hypotheses, (ii) hypothesis-free exome-wide, (iii) or genome-wide association analysis, (iv) and multivariate approaches such as clustering methods or machine learning to identify associations of genetic variants with outcomes of interest. We will perform meta-analyses using combined cohorts of discovery and replication datasets and previous studies reporting on the same genetic variants, where possible.

We will mainly use the software packages Stata (Stata Corporation, Austin, Texas), R (R Foundation for Statistical Computing, Vienna, Austria), and PLINK[51] for analyses. Depending on the sub-study and data availability, we will use different pipelines for quality control, filtering, and annotation.[52] Statistical significance tests will be two-sided and appropriate significance levels will be applied, adjusting for multiple testing where appropriate taking into account clinical co-variates, where possible (Bonferroni method, False Discovery Rate by Benjamini and Hochberg, or similar). Statistical uncertainty of estimates will be expressed as 95% confidence intervals.

#### 3.10. Power calculation

For each sub-study, we will calculate the power of the planned association analysis using appropriate tools, e.g. the Genetic Association Study (GAS) Power Calculator (https://csg.sph.umich.edu/abecasis/gas_power_calculator).[53] We will perform power calculations that are appropriate for the intended analyses such as genome-wide, exome-wide, or candidate-gene association studies. We will use for the different approaches the expected number of variants after filtering and define the adjusted cut-off p-value appropriate for multiple testing. We will estimate the sample size taking into account the outcome of interest incidence, the expected relative risk for possible risk variants, and minor allele frequency cutoff values using different models (dominant, additive, recessive, where appropriate).

#### 3.11. *In silico* and *in vitro* analyses

We will use computational *(in silico)* tools to estimate the effect of specific gene variants on gene regulation, splicing and expression of proteins (e.g. PolyPhen, SIFT, Human Splicing Finder, Matinspector).[54–57] We will choose *in silico* methods to identify genetic variants associated with complex disease mechanisms. Examples of such models are clustering methods including similarity network fusion[38] and PEGASUS[58]) and deep learning methods to explore interactions between genes and outcomes of interest.[59] We will use multiple outcomes in combination with multiple treatment exposures in suitable models to test their association with genetic variant data and identify genetic variants associated with multiple complications.[60, 61]

We will perform *in vitro* analyses using cell culture models relevant to the outcomes of interest. We will treat cell lines with antineoplastic agents or irradiate them and then perform transcriptomic analyses to identify genes that are differentially expressed after exposure to specific treatment modalities. We will utilize adapted approaches depending on the outcomes of interest to prioritize genes for further use in genotype-phenotype association studies. Through differential analysis of change in gene expression after treatment exposure, we will seek to identify candidate genes for further use in association studies. In hypothesis-free analysis methods (WGS or WES) we will perform *in vitro* studies to clarify the biological function of identified genes and genetic variants.[54, 55]

#### 3.12. Validation and replication

We will seek to validate variants identified by next-generation sequencing that were associated with the respective outcome of interest using a different method (e.g. Sanger sequencing or real-time PCR). After successful validation, we will seek to proceed to replication of identified variants in a targeted analysis of an independent cohort containing similar outcome information as the one analyzed in the primary dataset.

### 4. Discussion

The GECCOS study will enable genotype-phenotype association studies focusing on various health conditions in childhood cancer patients and survivors. The large inter-individual variability in response to antineoplastic treatments and occurrence of early and late complications is currently addressed mainly in a trial-and-error approach i.e. by delaying and adjusting treatment after occurrence of complications. Follow-up care is stratified by treatment exposure but not by genetic predictors.[62] The advantage of germline genetic risk variants is that this knowledge can be assessed when the workup of the neoplasm is made and then used early in the course of the treatment as they do not change over time. Such knowledge would allow to personalize treatment and follow-up care for individual patients before clinical signs of complications are present. Knowledge of genetic variants associated with treatment response will help maximizing treatment effect while reducing the risk for complications and finding the balance of treatment intensity in the light of increasing survival in childhood cancers. Genetic predictors will improve individual counselling of patients and their families and help developing individualized follow-up guidelines.

Assessing multiple outcomes taking into account multiple covariates including treatment exposures will help identify particularly vulnerable patients. As identified in previous research, many patients suffer from several complications after childhood cancer treatment.[4] Finding genetic variants associated with increased risk for multiple health conditions will help identifying gene variants that contribute to several organ system complications. This approach might help identifying patients which could most benefit from treatment adaptation and preventive measures to reduce complications.

GECCOS provides a legal and organizational platform on how to use sensitive genetic data with clinical information in association studies. It establishes structures that can be used by researchers for national and international collaborative studies. Germline genetic sequencing data generated in the GECCOS sub-studies will be stored after completion in BISKIDS hosted within the BaHOP biobank, Geneva University Hospital. Clinical data will remain in the described databases and only be temporarily linked for research studies increasing data safety. The populations of interest will overlap between sub-studies and sequenced datasets generated from participants included in completed sub-studies will contribute with their germline genetic data to subsequent studies. This growing resource will reduce costs for future studies, where only DNA of a fraction of the participants will have to be sequenced. We will favor whole genome and whole exome sequencing to create datasets that are not restricted to a specific research question but can be used for further research. We will then be able to address different questions with the same datasets. A further strength of GECCOS is the availability of a large clinical dataset collected since 1976, curated and updated follow-up information, and survival data regularly.

GECCOS will be limited due to the fact that Switzerland is a small country with a limited number of possible participants, which we can recruit for our research despite the nationwide and population-based sampling. This will be counteracted by international collaborations. Another issue with research on childhood cancer complications is that these health conditions are complex diseases with likely many mechanisms leading to a specific outcome. This makes identification of specific gene variants difficult. Many findings from studies were not replicated in independent datasets. Candidate-gene studies have particularly suffered from this. We will also explore novel methods to cluster and associate gene variants with clinical outcomes.

Our workflow combining a large dataset of clinical information with germline genetic data will enable genetic research on patient populations within Switzerland and facilitate collaborations with other research groups. As all childhood cancers are rare diseases by definition of the world health organization (WHO) with less than 1 in 2,000 people being affected, patient numbers are generally small. Research on rare childhood cancer subtypes or specific rare outcomes is only possible through international collaborations. We will provide a platform for these collaborations with the GECCOS study.

## Supporting information

Supplementary File: SPIRIT checklist

## Data Availability

The datasets that will be generated within GECCOS and/or analyzed during the study will not be publicly available due to concerns for anonymity of patients with childhood cancer being a rare disease. Datasets are available from the corresponding author on reasonable request for specific projects.

## Abbreviations table

ALL: Acute lymphoblastic leukemia
BaHOP: Biobank for Hematology and Oncology in Pediatrics
BISKIDS: Germline DNA Biobank Switzerland for Childhood Cancer and Blood Disorders
CNS: Central nervous system
CPS: Cancer predisposition syndrome
DNA: Deoxyribonucleic acid
EWAS: Exome-wide association study
GECCOS: Genetic risks for childhood cancer complications Switzerland GWAS Genome-wide association study
HSCT: Hematopoietic stem cell transplantation
IARC: International Agency for Research on Cancer
ICCC-3: International Classification of Childhood Cancer, third edition ISPM Institute of Social and Preventive Medicine
LCH: Langerhans cell histiocytosis
SCCR: Swiss Childhood Cancer Registry
SCCSS: Swiss Childhood Cancer Survivors Study
SPN: Second primary neoplasm
WHO: World Health Organization

## Declarations

### Ethics approval and consent to participate

The Geneva Cantonal Commission for Research Ethics has approved the GECCOS study (approval 2020-01723), and the BaHOP biobank (approval PB_2017-00533).

The **sponsor** of the GECCOS study is the University Hospital Geneva, Rue Willy-Donzé 6, 1211 Geneva 4, Switzerland; responsible person: Prof. Klara Posfay Barbe

The current **version** of the GECCOS protocol when this manuscript was submitted: Version 1.0; May 28^th^, 2020. Changes in the protocol will be communicated to collaborators by the principal investigator and the main study team, if relevant, and amendments made on clinicaltrials.gov.

## Consent for publication

Not applicable.

## Competing interests

The authors declare that they have no competing interests

## Funding

This study is supported by the CANSEARCH Foundation for BISKIDS, the host biobank BaHOP, the research study GECCOS, and salary support to Nicolas Waespe. Further funding comes from the Swiss National Science Foundation (31BL30_185396), and Swiss Cancer Research (KFS-4722-02-2019, KLS/KFS-4825-01-2019).

The funding bodies have no role in the design of the study and collection, analysis, and interpretation of data and in writing this manuscript.

## Authors’ contributions

NW: Conceptualization, Design, Methodology, Writing -all stages, Visualization.

SSt: Methodology, Design, Writing -Reviewing and Editing.

TN, CRSU: Methodology, Writing -Reviewing and Editing. DM, VM: Design, Writing -Reviewing and Editing.

MO, FGP, AOB, FB, LM: Writing -Reviewing and Editing.

CEK, AS: Design, Methodology, Writing -Reviewing and Editing.

MA: Supervision, Conceptualization, Design, Methodology, Writing -Reviewing and Editing.

All authors have approved the submitted version and have agreed both to be personally accountable for the author’s own contributions and to ensure that questions related to the accuracy or integrity of any part of the work, even ones in which the author was not personally involved, are appropriately investigated, resolved, and the resolution documented in the literature.

## Acknowledgements

We thank all childhood cancer patients, survivors, and families for participating in our study. We thank the study team of the Childhood Cancer Research Group, Institute of Social and Preventive Medicine, University of Bern, and SCCSS (Christina Schindera, Tomas Slama), the data managers of the Swiss pediatric oncology clinics (Claudia Althaus, Nadine Assbichler, Pamela Balestra, Heike Baumeler, Nadine Beusch, Sarah Blanc, Pierluigi Brazzola, Susann Drerup, Janine Garibay, Franziska Hochreutener, Monika Imbach, Friedgard Julmy, Eléna Lemmel, Heike Markiewicz, Annette Reinberg, Renate Siegenthaler, Astrid Schiltknecht, Beate Schwenke, and Verena Stahel) and the data managers and administrative staff of the SCCR (Meltem Altun, Erika Brantschen, Katharina Flandera, Elisabeth Kiraly, Verena Pfeiffer, Julia Ruppel, Ursina Roder, and Nadine Lötscher). We thank the study team of the Research Platform for Pediatric Oncology and Hematology at the Geneva medical school (Khalil Ben Hassine, Simona Jurkovic Mlakar, Fanny Muet, Mary Khoshbeen-Boudal, Laurence Lesne, Vid Mlakar, Shannon Robin, Yoann Sarmiento) and the Onco-Hematology Unit of the HUG (Fanette Bernard, Laurent Cimasoni, Violaine Guignon, Nelly Hafner-Bénichou, Rodolfo Lo Piccolo).

## Notes

### Competing Interest Statement

The authors have declared no competing interest.

### Clinical Trial

NCT04702321

### Author Declarations

Ethics approval and consent to participate The Geneva Cantonal Commission for Research Ethics has approved the GECCOS study (approval 2020-01723), and the BaHOP biobank (approval PB_2017-00533). The sponsor of the GECCOS study is the University Hospital Geneva, Rue Willy-Donze 6, 1211 Geneva 4, Switzerland; responsible person: Prof. Klara Posfay Barbe, The current version of the GECCOS protocol when this manuscript was submitted: Version 1.0; May 28th, 2020. Changes in the protocol will be communicated to collaborators by the principal investigator and the main study team, if relevant, and amendments made on clinicaltrials.gov.

