## Supplementary Tables for "Cohort-based association study of germline genetic variants with acute and chronic health complications of childhood cancer and its treatment: Genetic risks for childhood cancer complications Switzerland (GECCOS) study protocol"

**Supplementary Table 1:** In- and exclusion criteria for the established sub-projects on pulmonary complications, hearing loss, and second primary neoplasms.

| Outcome of interest | Inclusion criteria | Exclusion criteria |
|---|---|---|
| **Pulmonary dysfunction** | 1) **Irradiation** of any dose potentially including the lungs:<br>a. Total body irradiation;<br>b. Total lung irradiation;<br>c. Radiation to the chest including the lungs;<br>d. Spinal irradiation;<br>e. Radiation to the upper abdomen or the neck.<br><br>2) **Chemotherapies** with known and suspected lung-toxic agents:<br>a. Bleomycin;<br>b. Busulfan;<br>c. Nitrosoureas (Carmustine, Lomustine);<br>d. High-dose Methotrexate. | **Surgery** to the lungs (except biopsies) |
| **Hearing loss** | 1) **Irradiation** to the head of 30 Gy or more<br><br>2) **Chemotherapies** with known hearing-toxic agents:<br>a. Cisplatin;<br>b. Carboplatin;<br>c. Oxaliplatin<br><br>3) Survivors of leukemia, CNS tumors, neuroblastoma, soft tissue sarcomas, and germ cell tumors who were not exposed to established ototoxic treatments (as defined above) but who are suspected to be at risk for hearing loss due to potential additional risk such as other drugs like aminoglycosides or loop diuretics. | 1) **Pre-existing hearing loss** before start of the cancer treatment<br><br>2) **Surgery** involving the ear which were associated with hearing loss. |

| | | |
|---|---|---|
| **Second primary neoplasms** | **1) Cases with second primary neoplasms**<br><br>**2) Matched control design:** matched by demographic, primary cancer diagnosis, and treatment factors, age at primary diagnosis, follow-up time, year of primary neoplasm treatment, and exposure to relevant treatments (e.g. chest radiation, or alkylating agents), where appropriate.<br><br>**3) Case-cohort design:** Random selection of a subcohort from all childhood cancer survivors and retrieval of the same information as needed for the cases. | Identified from medical records, follow-up reports to the SCCR, linkage with cantonal cancer registries, death records, and questionnaire information (as defined by IARC)[1] |

**Supplementary Table 2:** Clinical information collected for the three established sub-projects on pulmonary dysfunction, hearing loss and second primary neoplasms.

| | | |
|---|---|---|
| **Pulmonary dysfunction** | **Spirometry** | Forced vital capacity (FVC), Forced expiratory volume in 1 second (FEV1), Peak expiratory flow (PEF), Maximum expiratory flow (MEF (25-75) |
| | **Body plethysmography** | Total lung capacity (TLC), Vital Capacity (VC), Functional residual capacity (FRC), Residual volume (RV) |
| | **Carbon monoxide diffusion capacity corrected for hemoglobin (DLCO)** | mmol/min/kPa corrected for hemoglobin if available |
| | **Personal history and symptoms** | Self-reported questionnaire information on recurrent pneumonias, chronic cough, and risk factors for lung problems (smoking, etc.) |

| | | |
|---|---|---|
| **Hearing loss** | **Audiometry** | Bilateral pure tone audiometry with air and bone conduction spanning 125 Hz to 8,000 Hz (where available up to 16,000 Hz) |
| | **Video otoscopy** | Assessment of the tympanic membrane (where available) |
| | **Personal history and symptoms** | Self-reported questionnaire information on hearing loss, tinnitus, hearing aid use, and exposure to noise |
| **Second primary neoplasms** | **Age at diagnosis** | Years |
| | **Date of diagnosis** | Month/ year |
| | **Type of diagnosis** | ICCC3 code; ICDO3 morphology, topography, behavior code |
| | **Laterality** | Left/ right/ bilateral/ medial/ not applicable |
| | **Relapse date** | Month/ year |
| | **Relapse type** | Local/ distant/ systemic/ other |
| | **Relapse location** | Organ and morphology |
| | **Treatment information** | Cumulative doses of individual antineoplastic agents and radiotherapy |

Legend: IARC, International Agency for Research on Cancer; SCCR, Swiss Childhood Cancer Registry; Hz, Hertz
